# Evaluation of at-home methods for N95 filtering facepiece respirator decontamination

**DOI:** 10.1101/2021.01.05.20248590

**Authors:** T.X. Chen, A. Pinharanda, N. A. Steinemann, K. Yasuma-Mitobe, E. Lee, J. Hahn, L. Wu, S. Fanourakis, D. S. Peterka, E.M.C Hillman

## Abstract

N95 filtering facepiece respirators (FFRs) are essential for the protection of healthcare professionals and other high-risk groups against Coronavirus Disease of 2019 (COVID-19). In response to shortages in FFRs during the ongoing COVID-19 pandemic, the Food and Drug Administration issued an Emergency Use Authorization permitting FFR decontamination and reuse. However, although industrial decontamination services are available at some large institutions, FFR decontamination is not widely accessible.

To be effective, FFR decontamination must 1) inactivate the virus; 2) preserve FFR integrity, specifically fit and filtering capability; and 3) be non-toxic and safe. Here we identify and test at-home heat-based methods for FFR decontamination that meet these requirements using common household appliances. Our results identify potential protocols for simple and accessible FFR decontamination, while also highlighting unsuitable methods that may jeopardize FFR integrity.

**One sentence summary:** Survey of at-home methods for N95 respirator decontamination using heat and evaluation of their effects on N95 respirator integrity.

## INTRODUCTION

Coronavirus Disease of 2019 (COVID-19), caused by the severe acute respiratory syndrome coronavirus 2 (SARS-CoV-2), is predominantly transmitted through exposure to short-range droplets and aerosols^1^. To protect against infectious airborne particles^2^, filtering facepiece respirators (FFRs) such as N95 respirators^3^ are recommended by the United States Center for Disease Control and Prevention (CDC) for healthcare professionals ^4^. With appropriate donning and doffing procedures, combined with proper fit, N95 FFRs can effectively limit cross-contamination and the transmission of COVID-19 ^5^.

The global increase in demand for personal protective equipment (PPE) during the COVID-19 pandemic has resulted in FFR shortages ^3^. This led the Food and Drug Administration (FDA) to issue an emergency use authorization for N95 FFR reuse and decontamination during the pandemic ^6^. To decontaminate FFRs, a minimum 3-log (99.9%) reduction in SARS-CoV-2 viral activity ^7^ needs to be achieved. In addition to inactivating the virus, decontamination strategies must maintain overall FFR integrity, as measured by fit (ability to form a tight seal) and filtration (ability to limit penetration of particulates) ^8^.

Several decontamination methods have been shown to satisfy these criteria, including, ultraviolet (UV) germicidal irradiation (radiation-based); hydrogen peroxide vapor (HPV) (chemical-based); and heat-humidity treatments ^9^. Unfortunately, these procedures can be difficult to implement at a large scale, and/or present safety risks at a small scale, especially in resource-limited environments. HPV, for example, is a severe irritant that can be hazardous to the lungs and eyes. Most UV-C light sources have spectral variation, and emit wavelengths that can damage the skin or eyes. Proper decontamination by UV-C light requires every surface to receive a sufficient dose of radiant energy, which is challenging to achieve for FFRs with three-dimensional shapes and metal inserts ^10^.

Another significant obstacle is that methods that provide full sterilization using heat, such as autoclaving, are likely to damage FFR integrity. As a result, existing FFR decontamination methods, even though effective against SARS-CoV-2, do not typically eliminate all pathogens. To prevent cross-contamination from pathogens that haven’t been eliminated, decontaminated FFRs need to be returned to the original user, posing a major logistical challenge on an institutional scale, particularly during a healthcare crisis. Therefore, establishing safe and economical decontamination methods that can be implemented by the end user is both necessary and urgent.

In this study, we recognized the potential of widely available, commercial household heat-generating appliances for FFR decontamination. We evaluated the ability of several appliances to maintain decontamination conditions, and tested the effects of repeated treatments on FFR fit and material filtration performance. Of the devices tested, we identify rice cookers with thermostatically-controlled warming functions as the most reliable at maintaining conditions for heat-based decontamination while retaining FFR fit and filtration integrity for up to five treatment cycles. We confirmed that boiling respirators is damaging to FFR fit.

## RESULTS

### Establishing decontamination parameters

Previous studies suggest that temperatures of 70-85°C for 30 minutes at 50% humidity ^11,12^ or 60 minutes with dry heat (increased temperature alone without added moisture) ^13^ are sufficient to inactivate SARS-CoV-2 from FFR fabric, while also preserving fit and filtration efficiency for up to five rounds of treatment, in line with CDC’s recommendations ^9^. In contrast, treatment with steam autoclaves or immersion boiling should inactivate SARS-CoV-2 from FFRs, but is likely to reduce FFR filtration efficiency and impact fit after as few as one round of treatment ^14–16^ (Table S1).

### Identifying home appliances capable of accurately attaining decontamination conditions

After considering a wide range of commercial heat-generating appliances we excluded those with (1) minimum temperatures above 70ºC, such as standard gas ovens ^21^, (2) maximum temperatures below 70°C, such as commercial tumble dryers and hair dryers^22^ and (3) appliances that pose potential safety hazards, including microwaves which ^17,18^ may cause FFRs with metallic components to spark or the plastic in the straps to melt ^19,20^. Based on these temperature and safety criteria, we selected rice cookers with *Keep Warm* settings and sous vide (immersion cooker) machines for further evaluation ^21,22^. Two types of rice cookers with *Keep Warm* settings - the Cuckoo (CR-0655F, retail ∼$110), and the Aroma (ARC-743-1NGB, retail ∼$20) - and an AuAg Immersion Circulator sous vide machine (A808, 950 W, retail ∼$50) were selected for testing.

### Assessing basic device performance

Initial testing was performed to confirm whether each device could reach and maintain the target temperature of 70ºC for one hour (Table S2). The *Keep Warm* setting of the Cuckoo rice cooker (71.7 ºC ± 0.3 ºC) and the AuAg Immersion Circulator sous vide machine (70.1 ºC ± 0.7 ºC) were confirmed to have precise temperature control. In contrast, the *Keep Warm* setting of the Aroma rice cooker does not allow for precise temperature control, with recorded temperatures of 67.5ºC ± 7.8 ºC across the hour-long treatment, this appliance was excluded from all subsequent tests. This result highlights the importance of confirming each device’s ability to maintain the specified temperature range before use.

We also tested for spatial variations in temperature in the Cuckoo rice cooker by measuring the temperature at one central and two peripheral locations of an untreated test FFR (Table S2, Fig S1). Confirming an even distribution across the FFR during the treatment cycle, the temperature at all three FFR locations stabilized close to 70ºC after the initial heating period (70.5ºC ± 0.4ºC at both lateral locations and 71.7ºC ± 0.3ºC at the central location).

### Evaluating the effects of appliance-based heat treatment on quantitative FFR fit

We tested the effects of three different decontamination protocols on FFR fit and function: 1) boiling at 100ºC for 10 minutes, 2) warming to 70ºC for 60 minutes using the Cuckoo rice cooker and 3) warming to 70ºC for 60 minutes using the sous vide machine (Fig 1). Even though boiling was expected to impair FFR function, it was tested as it would be particularly simple to perform at home ^8,15^.

**Fig 1.**
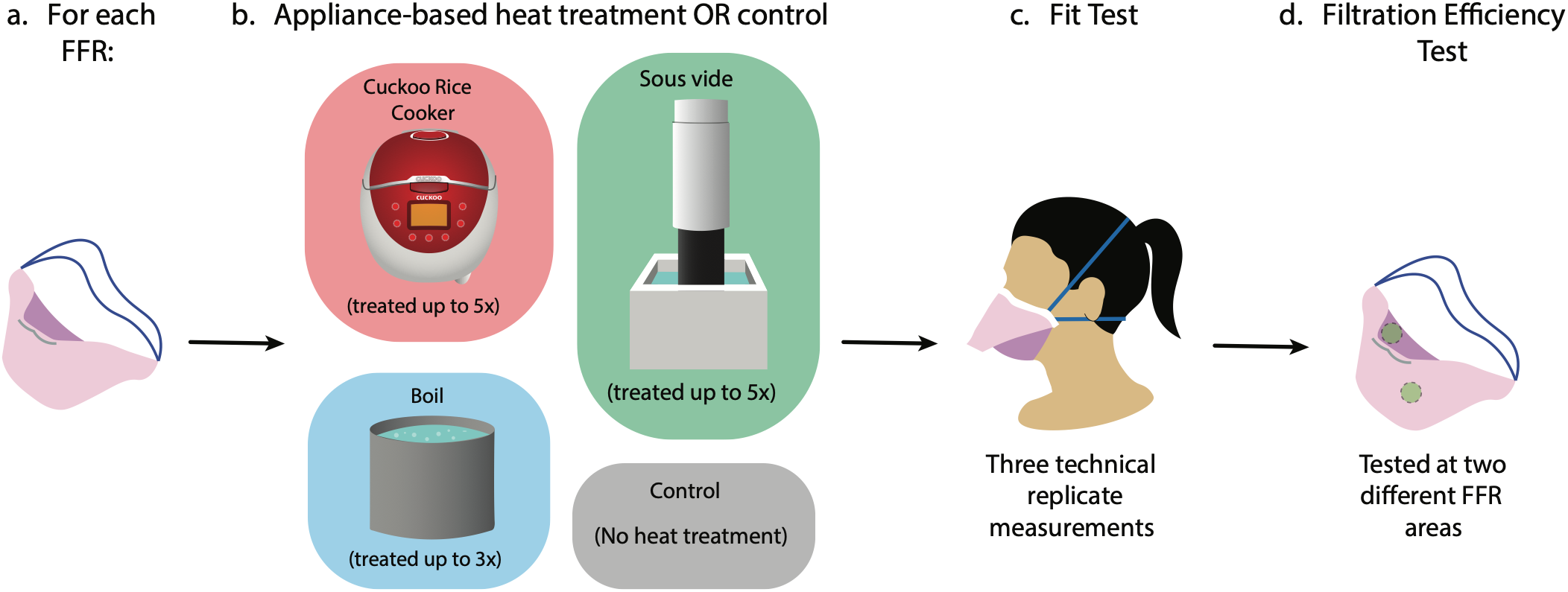
Experimental workflow for Halyard Fluidshield 3 (TC-84A-7521) FFRs. **(a)** Each FFR was blindly assigned to either **(b)** *x* round(s) of appliance-based heat treatment or to a control group. **(c)** Treatment-exposed and control FFRs were fit tested using the built-in N95 setting of a TSI PortaCount Pro+ Respirator Fit Tester 8038. **(d)** The effect of appliance-based heat treatment on FFR filtration performance was evaluated for treatment and control FFRs using a modified version of the standard NIOSH test.

Experiments were performed on two models of donated NIOSH-approved FFRs: the Halyard Fluidshield 3 (TC-84A-7521) (Fig S2a) and the Uniair San Huei SH3500 (TC-84A-4313) (Fig S2b). The Halyard Fluidshield 3 filter facepiece respirator was included because it is a commonly used respirator at major hospitals. The Uniair San Huei was among the few FFRs that could be procured when there were severe FFR shortages, and was chosen for comparison to the Halyard FFR. This research was conducted when PPE supplies were limited so care was taken to minimize the total number of FFRs used in these experiments (Fig. 1 and Supplemental Methods for full details of treatments and fit testing protocols).

Individual FFRs underwent one to five rounds of heat treatment followed by quantitative fit testing and material filtration assessment (Fig 1b). Control FFRs were not exposed to any treatment (Fig 2a). FFRs assigned to the rice cooker were placed in breathable paper bags to mimic how a home user would be advised to handle contaminated FFRs (Fig 2f) ^9^. For heat treatment with the sous vide machine, FFRs were sealed in a polypropylene plastic bag before being placed in water and two 140 g weights were placed on top of the plastic bag to ensure full submersion (Fig 2g). For boiling, the fabric was immersed directly in water while the elastic straps were kept above the water line to prevent altering their elasticity (Fig 2b) in accordance with a previous study ^8^. For all three conditions, extra care was taken to avoid deforming the FFR.

**Fig 2.**
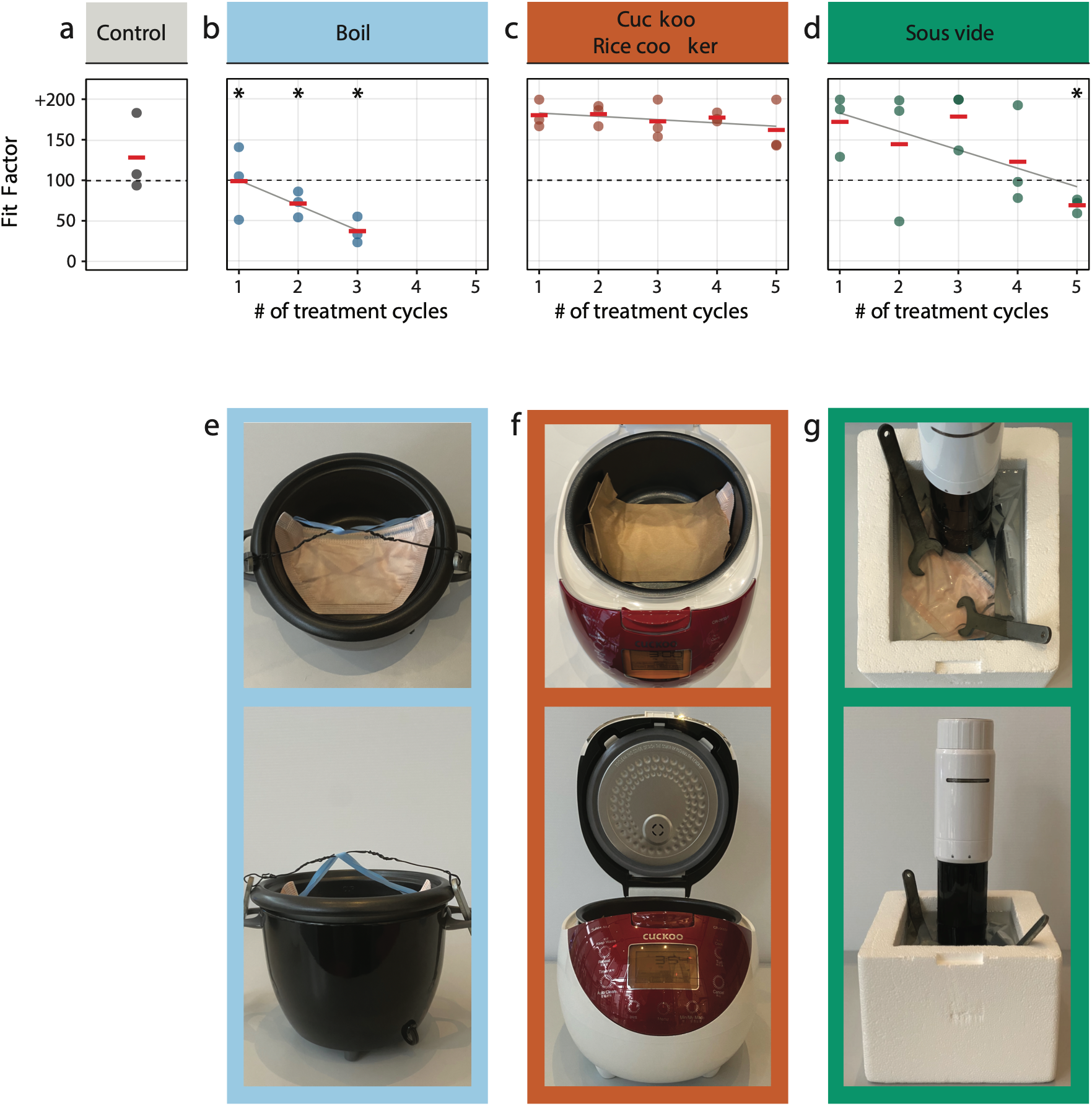
Average Halyard Fluidshield 3 FFR fit factor before and after treatment. Fit factor as a function of treatment cycles for each condition. Photograph of the heat generation appliance shown in the panel below their respective fit factor profile: **(a)** control, **(b** and **e)** boiling, using **(c** and **f)** a rice cooker and **(d** and **g)** a sous vide machine. Fit factor measures were repeated three times for each FFR (individual dots) and average fit factor is reported to the nearest integer (red bar). Grey regression lines quantify fit factor as a function of the number of treatment cycles. Asterisks indicate significant differences in fit between treatment group and control (ANOVA, p < 0.05).

Treatment-exposed and control FFRs were fit tested using the built-in N95 setting of a TSI PortaCount Pro+ Respirator Fit Tester 8038. Quantitative fit of FFRs is tested for each individual user in seven standardized test conditions: *normal breathing, deep breathing, head side to side, head up and down, talking, bending over*, and *normal breathing*. These results are commonly combined into an overall fit factor computed as the geometric mean of the individual conditions. FFRs with a mean fit factor of 100 or greater, pass the fit test. Three technical replicates of all measure were taken for each FFR (Fig 2 a-d).

As shown in Fig 2c, exposing Halyard Fluidshield 3 FFRs to 70**°**C of dry heat for 60 minutes in the Cuckoo rice cooker up to five times did not decrease their overall fit quality significantly (F(1, 4) = 2.57, p = 0.18). Treatment with the sous vide machine and boiling, on the other hand, significantly decreased fit quality of Halyard Fluidshield 3 FFRs, leading to failed fit testing after five (F(1, 4) = 36.76, p < 0.01; Fig 2d) and one (F(1, 4) = 12.54, p < 0.05; Fig 2b) cycles of treatment, respectively (Table S3). We conclude that, among the three tested methods, the Cuckoo rice cooker, and by extension, dry heat, is best at preserving Halyard Fluidshield 3 FFR fit.

The overall fit of Uniair FFRs on our pool of testers was found to be poor, and even the control FFRs failed two out of three technical replicate fit test measurements (Fig S3, Fig S4, Table S4 and Table S5). Based on the poor fit performance and lack of trends in fit quality with treatment conditions (Fig S4), we cannot draw meaningful conclusions on how the number of treatment repetitions affected the Uniair FFR fit. These results underscore the importance of performing regular quantitative fit testing of FRRs to each individual user to ensure proper fit under a respiratory protection plan ^23^. While it is possible that trends in fit following heat-treatment hold for similarly manufactured FFRs, the conclusions of this study regarding FFR fit after heat treatment thus pertain primarily to the Halyard Fluidshield 3 (TC-84A-7521).

### Comparison of individual fit test categories gives insights into how to improve fit

Quantitative fit testing of FFRs involves a series of seven test conditions (*normal breathing, deep breathing, head side to side, head up and down, talking, bending over*, and *normal breathing*), but results are often only examined using the mean fit factor. However, a more detailed examination of results for each part of the test can provide valuable insights (Fig 3).

**Fig 3.**
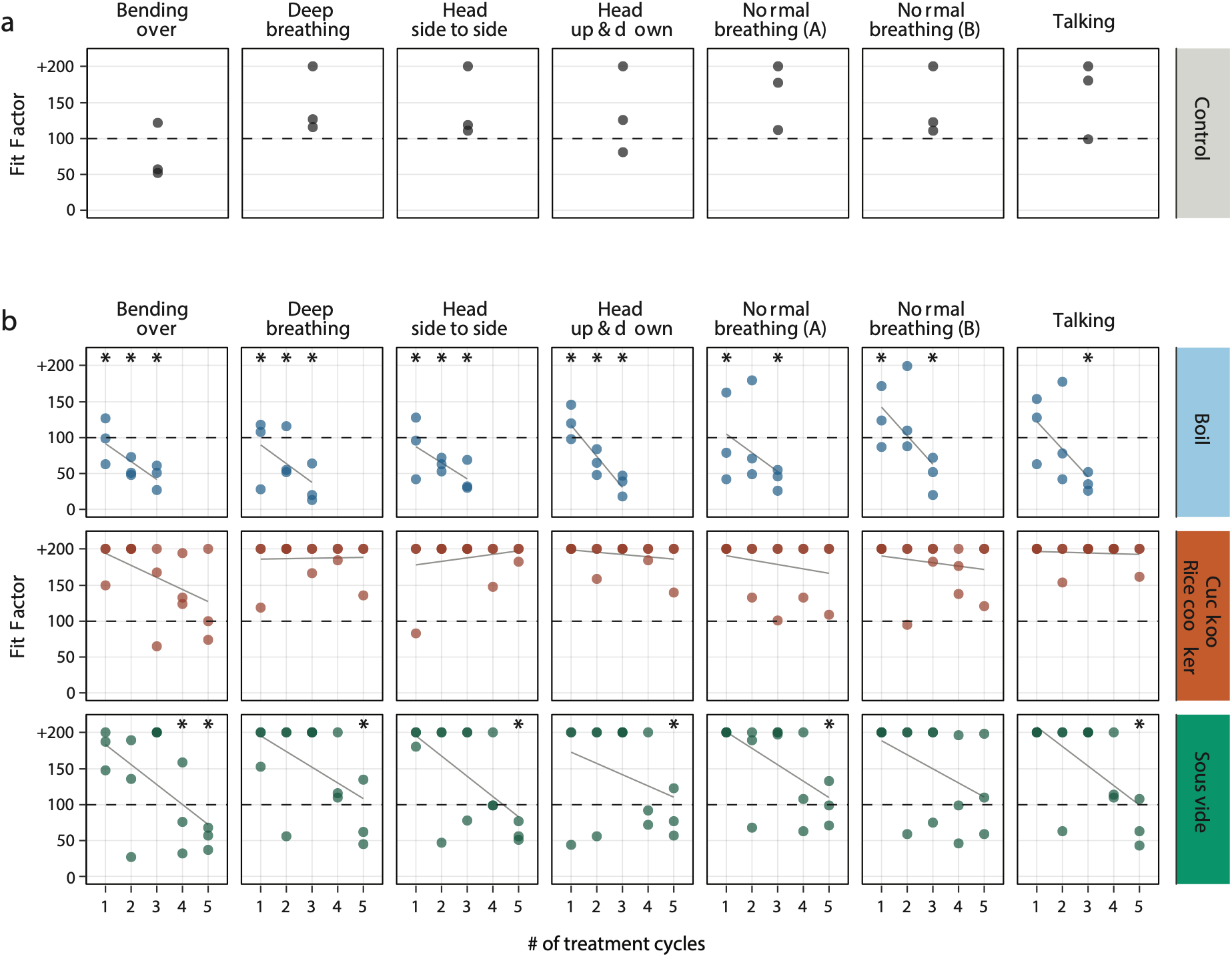
Halyard Fluidshield 3 FFR fit for seven standardized testing conditions. The fit of control **(a)** and heat-treated **(b)** FFRs was assessed three times in each of seven testing conditions (individual dots) for each heat-treatment and control group. Grey lines in (b) signify linear regression fit of fit factor over treatment cycle (Table S7 and S8). Asterisks indicate significant differences in mean fit between treatment group and control (ANOVA, p < 0.05).

FFRs treated in the Cuckoo rice cooker passed the overall fit test after five treatment cycles, however, there was a progressive decrease in the goodness of fit when the subject was *bending over* (Fig 3, Table S6). Boiling and heat treatment using a sous vide machine progressively decreased fit scores in all conditions. For sous vide machine treatment, while FFRs still passed the overall fit test after four rounds of treatment (FF = 123), fit factors while *bending over* were significantly lower than in untreated FFRs (p < 0.05) (for further details see Fig 3, Table S6, Table S7, Table S8).

Our results suggest that rice cookers and sous vide machines are viable options for the decontamination of N95 FFRs. Moreover, they highlight the 1) importance to make technical replicate measurements, which is currently not required for fit testing, and 2) record each category’s fit factor. While dry heat is the method that preserves fit best, we propose that paying special attention to movement-specific fit testing results when fitting a respirator could aid the end user to adjust their behavior while wearing FFRs (e.g. kneeling vs. bending).

### Evaluating the effects of appliance-based heat treatment on FFR filtration performance

The effect of appliance-based heat treatment on FFR filtration performance was evaluated for treatment and control FFRs that had passed at least one round of treatment (FF ≥ 100, Uniair FFRs were excluded). We developed a modified version of the standard NIOSH test (TEB-APR-STP-0059) to assess the penetration of non-neutralized saline aerosols as the difference in particle counts before and after passing an aerosol through the FFR material (Fig 4a). FE was measured for particles in six different size classes (0.3 μm, 0.5 μm, 1 μm, 2.5 μm, 5 μm, and 10 μm) by a calibrated particle counter. Like similar custom-built devices ^22^, our apparatus was designed to normalize for constant flow rates regardless of filter impedance (Fig 4a, Table S9). Furthermore, our experiments were conducted with a steady flow rate per unit area of the filter that exceeds the rates expected under standard NIOSH testing conditions, which generally decreases filtering efficiency compared to lower flow rates ^24–26^, suggesting a conservative estimate of FE. Testing was performed on two different central sites on each heat-treated and control FFR.

**Fig 4.**
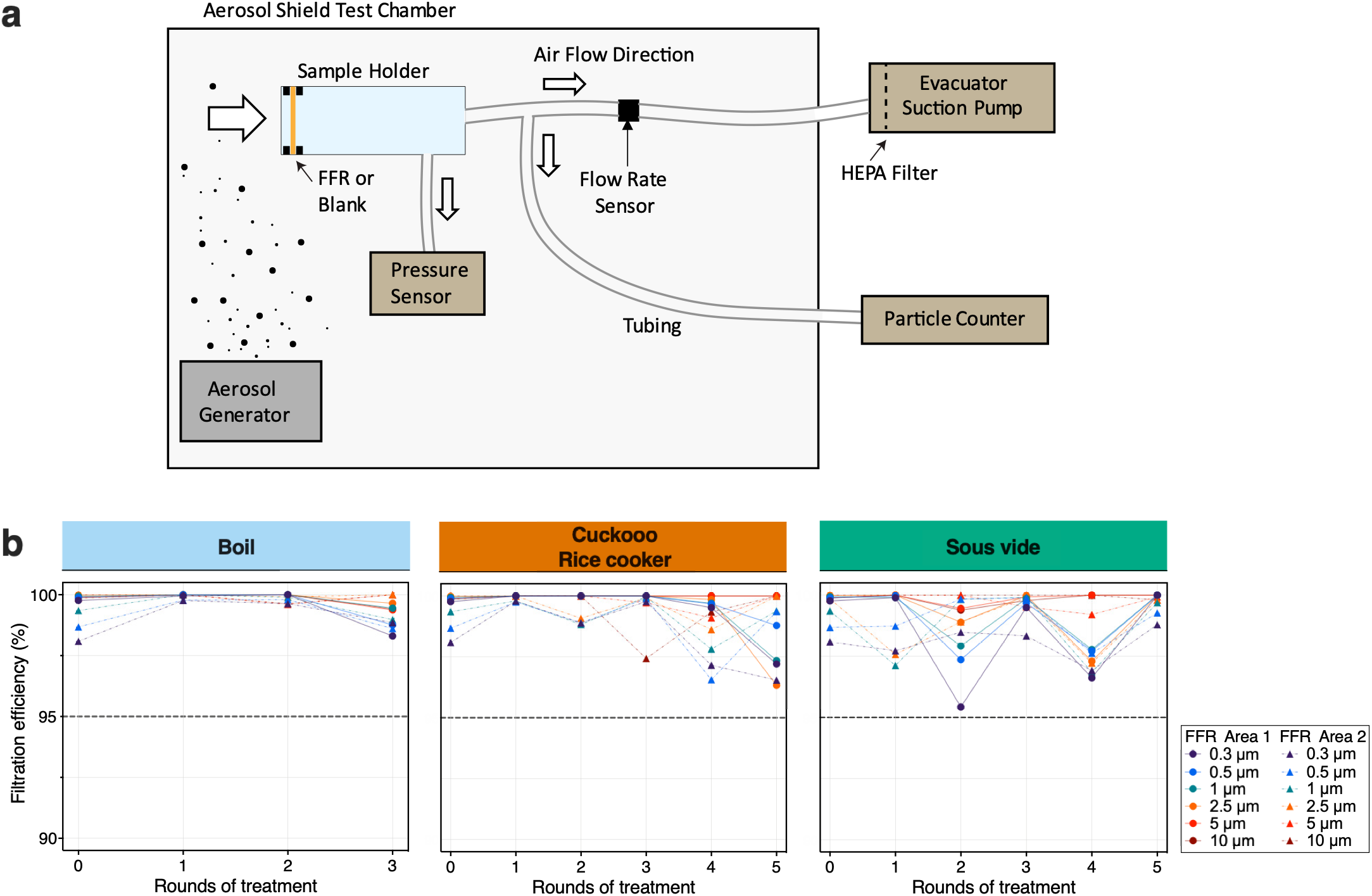
Filtration efficiency for control and heat-treated Halyard Fluidshield 3 FFRs. **(a)** Filtration efficiency test experimental setup. A humidifier confined by an aerosol shield test chamber was used as an aerosol generator. FFRs were attached to a sample holder and air and any suspended particles were drawn through the FFR by an evacuator suction pump and diverted to a particle counter. FFR impedance was measured through a pressure sensor attached to the sample holder. **(b)** The filtration efficiency for six different particle sizes and for two regions of heat-treated and control FFRs. None of the treatment methods reduced Halyard Fluidshield 3 FFR filtration performance below 95% for particles of any size as shown in Table S9.

Comparisons between filtration efficiencies in the control and heat-treated FFRs indicate that none of the tested decontamination strategies reduced FFR FE below the 95% required of ‘N-95’ FFRs (Fig 4b, Table S9). To both validate the sensitivity of our custom-built filtration test rig and to demonstrate how even small leaks can significantly affect effective filter performance, we used an 18-gauge needle to introduce small holes in the filter after normal testing. FE decreased as a function of the number of pinholes introduced into the FFR material (Fig S5), and pressure across the FFR material decreased at a flow rate of 10 lpm (Table S10). The validity of these results is further supported by passing scores during the quantitative fit tests, as any damage to the N95 FFR material would compromise the overall filterability of the respirator and result in the FFR failing the fit test. On the other hand, FE testing also demonstrates that flow and filtration properties of the FFR material are not the only important consideration, as proper fit is critical to maintain a high level of protection.

## DISCUSSION

It has been previously shown that SARS-CoV-2 can be inactivated in FFRs by one-hour treatments with dry 70**°**C dry heat^13^. Under these conditions, some FFRs have been shown to preserve fit and filtration efficiency for up to ten decontamination cycles ^13,27^. Here we assessed three heat-based methods that are likely to inactivate SARS-CoV-2 using common household appliances and simple methods: a rice cooker, a sous vide machine and boiling in water.

We compared the fit and filtration properties of two FFR models, the Halyard Fluidshield 3 and the Uniair San Huei, after up to five sequential rounds of heat treatment to those of untreated FFRs. Even before treatment, the Uniair San Huei FFR exhibited widely varying fit test results in our pool of testers, and was thus excluded from further analyses. This result emphasizes the importance of fitting FFRs to each individual user, as proper fit is paramount for appropriate protection.

We show that the Halyard Fluidshield 3 fit and filtration efficiency are preserved for up to five rounds of treatment using the *Keep warm* function of a rice cooker with precise temperature control. As the CDC does not recommend re-using FFRs more than five times, we did not subject the respirators to more than five rounds of treatment ^9^. Sous vide machine treated FFRs preserved fit only up to four rounds, possibly because the respirator was slightly deformed by the water and weights. FFRs boiled in water failed the fit test after just one round of treatment.

By examining fit test results in more detail, we found that for all decontamination strategies the fit factor, and therefore protection, was reduced the most and after the fewest treatment cycles while *bending over* (Fig 3). This indicates that a poor fit under this condition may serve as an early warning sign for a decrease in overall fit.

Flow and filtration analysis of treated FFRs demonstrated that none of the heat treatment methods lowered filtration efficiency below the required 95%. Interestingly, even boiling for up to three treatment cycles did not lower the FE of the Halyard Fluidshield 3 FFRs below this threshold. Although this observation appears to be in conflict with published results, those experiments tested different FFR models^28^. The Halyard Fluidshield 3 was designed to protect against fluid splashes in the operating room and is considered to be water-resistant. Further research is warranted to determine whether splash-protective respirators, in general, are more resistant to changes in FE during humid or water-immersive treatment conditions than conventional FFRs^8,27^, although it is important to note that we saw significant degradation in fit and thus efficacy with boiling, making it an unsuitable approach for decontamination overall.

While not recommended for clinical or routine use, the decontamination methods evaluated here demonstrate that household appliances could constitute a low-cost strategy for N95 decontamination that could be performed in resource-constrained settings or situations where no viable alternative exists. Before attempting decontamination, however, it is essential for the user to verify that the selected appliance can stably maintain a temperature of 70°C for 60 minutes with spatial homogeneity across the device. Additionally, after each decontamination cycle, FFRs need to be examined for damage, burns or tears. As highlighted by differences in the goodness of fit between FFR models and between individual FFRs exposed to the same decontamination treatment, it is further imperative to carefully examine FFR fit before the first use and after each treatment cycle to ensure tight fit of the FFR all around the face.

It is also important to note that exposure to dry heat at 70°C does not result in sterilization, the complete elimination of all forms of microbial life. Pathogens such as *Clostridium difficile* will not be eliminated by treatment methods like the ones described in this study, and thus, treated FFRs should only be re-used by the same individual^29^.

## MATERIALS AND METHODS

The Columbia University Institutional Review Board reviewed this study’s protocol and determined that ethical approval could be waived (IRB reference AAAT5503). All necessary participant consent was obtained.

### Assessing basic device performance

Initial assessment of whether each device could reach and maintain the target temperature of 70ºC for the treatment duration of one hour was performed. For each rice cooker, a temperature sensor was placed in an elevated plastic tub containing 120 ml of water within the inner pot while the rice cookers were heated using the *Keep Warm* setting (Table S2). For the sous vide machine, a temperature sensor was immersed in a polystyrene box with 3.2 L of water and warmed using a sous-vide temperature setting of 70ºC. All device temperatures were recorded during initial heating and then for one hour after each appliance’s temperature had stabilized. The total process time, including time to reach the target temperature, were also recorded (Table S2, Fig S1).

To confirm that the temperatures were distributed evenly across the FFR during the treatment cycle (70ºC for 60 minutes), we repeated these measures in the Cuckoo rice cooker at one central and two peripheral locations of an untreated test FFR (Table S2, Fig S1). For the Cuckoo rice cooker, the temperature at all three FFR locations stabilized close to 70ºC after the initial heating period (70.5ºC ± 0.4ºC at both lateral locations and 71.7ºC ± 0.3ºC at the central location).

### FFR treatment

We tested the effects of three different decontamination protocols on FFR fit and function: 1) boiling at 100ºC for 10 minutes, 2) 70ºC for 60 minutes using a Cuckoo rice cooker and 3) 70ºC for 60 minutes using a sous vide machine (Fig 1). Tests were performed on two NIOSH-approved N95 FFR models, the standard Halyard Fluidshield 3 (TC-84A-7521) and the Uniair San Huei SH3500 (TC-84A-4313) (Fig S2). FFRs were randomly assigned to either one of the three heat-treatment or a control group. Both the experimenter and test subject were blind to this assignment. FFRs were subjected to up to three cycles of decontamination by boiling or up to five cycles of dry heat decontamination (Fig 1).

Temperature within each appliance used for dry heat treatment was recorded throughout the treatment with a wireless sensor (Inkbird Thermometer IBS-TH1) for the rice cooker and MeatStick Wireless Thermometer (4335995327) for the sous vide machine. Both thermometers were benchmarked against a calibrated thermometer (Oakton Acorn Series pH 5 Meter and Thermometer). For dry heat treatment in the rice cooker, FFRs were placed in breathable paper bags inside the preheated rice cooker (Fig 2e) after which the temperature was maintained at 70 ºC for 60 minutes. For sous vide treatment, FFRs were sealed in double protection Ziploc© Freezer Bags before being placed in water preheated to 70 ºC. In order to completely submerge the FFRs, two 140 g weights were placed on top of the plastic bag (Fig 2g). The temperature of the sous vide machine was set to 70 ºC for 60 minutes. FFRs subjected to boiling were submerged directly in 1.3 L of boiling water at 100 ºC within a 1.4 L pot (Fig 2c) and were drained and allowed to air-dry overnight post-treatment.

### FFR fit test

Quantitative fit tests were performed using the manufacturer provided “N95” setting of a calibrated TSI PortaCount Pro+ Respirator Fit Tester 8038 with ambient particulates and aqueous aerosols produced by an ultrasonic humidifier (New York city tap water). Adherent to the Occupational Safety and Health Administration’s (OSHA’s) Respiratory Protection Standard (29CFR 1910.134), the Condensation Nuclei Counter (CNC) Quantitative Fit Test for Filtering Facepiece Respirators was used ^23^. This test reports a quantitative fit factor score for sequential test conditions (*normal breathing, deep breathing, head side to side, head up and down, talking, bending over*, and *normal breathing*) as well as an overall fit factor computed as the mean of the scores achieved in the individual conditions. Based on the OSHA Respiratory Protection Standard, an overall fit factor of ≥ 100 is required for passing, regardless of performance on each test component ^23^. It should be noted that the system gives a maximum possible score of 200+ reported as 201 ^30^.

The CNC quantitative fit test was performed three times for each FFR. All tests were performed on the same test subject to control for variability in facial features. As per CDC recommendations, a user seal check was performed prior to each fit test measurement and adjustments were made until the user deemed that there were no detectable air leaks during forceful breathing ^9^.

### FFR filtration efficiency test

Due to the limited availability of NIOSH test apparatus, a modified version of the standard procedure (TEB-APR-STP-0059) was developed to determine the particulate filter efficiency of N95 FFRs ^31^. FFR fabric was challenged by a NaCl aerosol generated using a humidifier (Vicks© Filter-Free Cool Mist Humidifier) filled with 2% saline solution. Non-neutralized particles were released into a polycarbonate test chamber (∼150 L) with a minimum 0.3-μm particle count of 5000 particles per ft^3^. Each FFR was manually secured over an iso-KF 25 fitting. After ∼10 cm of ∼25 mm ID stainless steel tubing, another KF-25 to barbed fitting joined via 3/8” ID silicone tubing connected to the both the calibrated particle counter (Extech Instruments VCP300) and a high-performance surgical smoke removal device used as an evacuator (Buffalo Filter VisiClear) (Fig 4A). Pressure drop across the filter was monitored with a Sensirion SDP8-500 differential pressure sensor, and the air flow volume was monitored by a Sensirion Mass Flow Meter (part number SFM3300-D). Both devices were read through their I2C communications channel through a custom program running on an Arduino Uno and then recorded on a PC at rates over 10 Hz.

Tests were conducted at a flow rate of 10 lpm with a 4.54 cm^2^ area of effective filtration, which exceeds the rate of 85 lpm used during NIOSH testing when our filter patch is scaled proportionally to the area of the entire FFR ^32^. We compensated for any variation in the impedance of the FFR material by changing the strength of the suction of the evacuator to obtain a flow rate of 10 lpm through each FFR or blank prior to each filtration test.

Before measuring the filtration efficiency of each treated or control FFR, the baseline ambient particle count within the aerosol shield chamber was measured as the particle count downstream of an open nozzle. First, particles were suctioned through either a blank or FFR at a flow rate of 10 lpm for 20 seconds, then the suction was switched off. Following a 5 second pre-counting sample draw, the cumulative number of same-sized particles downstream of each FFR or blank were counted over a 20 second sampling period at a 2.83 lpm sampling rate controlled by the particle counter’s internal pump. The total volume of the internal tubing, sensors, and stainless port was ∼1.35 liters, and designed such that the bulk of the air sampled by the particle counter during its total 25 second counter operation was air drawn through the filter unit at the 10 lpm volume flow rate, not air drawn through the filter by the pump action of the particle counter. Particles binned into 0.3 μm, 0.5 μm, 1 μm, 2.5 μm, 5 μm, and 10 μm size classes were measured. The filtration efficiency (FE) was calculated using the equation,

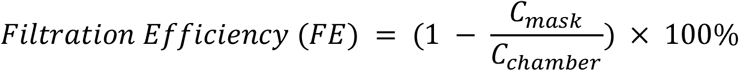

where *C*_*mask*_ and *C*_*chamber*_ denote the concentrations of particles downstream of the FFR and within the aerosol chamber, respectively. For each FFR, flow test measurements of two different central N95 material areas were obtained to sample the variance.

To assess the sensitivity of our modified flow test, two FFR samples were punctured with an 18-gauge needle up to three times (Fig S5, Table S10). Each puncture was at most 0.3% of the filter area, yet reduced filtering efficiency of the 0.3 μm particles by roughly 25% (e.g., from >95% to 75%). This large reduction in efficiency from such a small leak highlights the importance of proper fit, with no voids, for effective protection.

### Statistical analysis

All statistical analyses were run in the R software environment (v4.0.1). The decrease of fit factors over repeated treatment was evaluated using linear regressions (stat_smooth function, method=lm, ggplot2 v3.3.0). Variance component analyses were calculated with nested one-Way ANOVA (aov(score∼treatment+Error(FFR)). F-values and significance levels for the F-statistics reported ^33^.

## Data Availability

All data is available in the main text or the supplementary materials.

## Acknowledgments

We thank Richard Hormigo for circuit design for the filtration efficiency testing rig; David Brenner for helpful discussions; VisiClear for the Buffalo Filter VisiClear; Zuckerman Institute and Columbia Environmental Health and Safety for the space to perform experiments; Columbia Researchers Against COVID-19 for assembling this team; Wearing is Caring; Columbia University Institutional Review Board for assessing these experiments; Peter Andolfatto, Rui Costa, Ivaylo Ivanov, and Andrés Bendesky for support throughout the project; Anil Lalwani for providing the FFRs used in this study.

## Author contributions

T.X.C, A.P, D.S.P and E.M.C.H conceived the study and designed the experiments. T.X.C. and A.P. performed the experiments, analyzed the data and wrote the manuscript. N.A.S wrote the manuscript. K.Y-M. performed the literature survey. T.X.C, A.P, E.M.C.H and L.E. made the figures. D.S.P. designed and assembled the FFR filtration efficiency test and provided training. N.A.S, J.H. and L.W. provided specialist support. S.F. provided training on the PortaCountPro+. T.X.C, A.P, N.A.S and E.M.C.H wrote and applied for the IRB. All the authors commented and edited the manuscript.

## Data availability

All data is available in the main text or the supplementary materials.

