## Supplementary material for "Evaluation of at-home methods for N95 filtering facepiece respirator decontamination": Supplmentary_information

**Title:**

**One sentence summary:**

Survey of at-home methods for N95 respirator decontamination using heat and evaluation of their effects on N95 respirator integrity.


**Authors:**

Chen, T.X. [1]*, Pinharanda, A. [2]*, Steinemann, N. A. [1], Yasuma-Mitobe, K. [3], Lee, E. [4], Hahn J. [5], Wu L. [4], Fanourakis, S. [6], Peterka, D. S. [1]†, Hillman, E.M.C [1,5,7]†

* These authors contributed equally to this work

**Affiliations:**

[1] Mortimer B. Zuckerman Mind Brain Behavior Institute, New York, NY, 10027, USA

[2] Department of Biological Sciences, Columbia University, New York, NY 10027, USA

[3] Department of Microbiology and Immunology, Columbia University, New York, NY 10032, USA

[4] Columbia College, Columbia University, New York, NY 10032, USA

[5] Department of Biomedical Engineering, Columbia University, New York, NY 10027, USA

[6] Environmental Health and Safety, Columbia University, New York, NY 10032, USA

[7] Department of Radiology, Columbia University, New York, NY 10027, USA

**LIST OF SUPPLEMENTARY MATERIALS:**

- Supplementary Methods
- Supplementary Figs S1 – S5
- Supplementary Tables S1 – S10

**SUPPLEMENTARY FIGURES:**

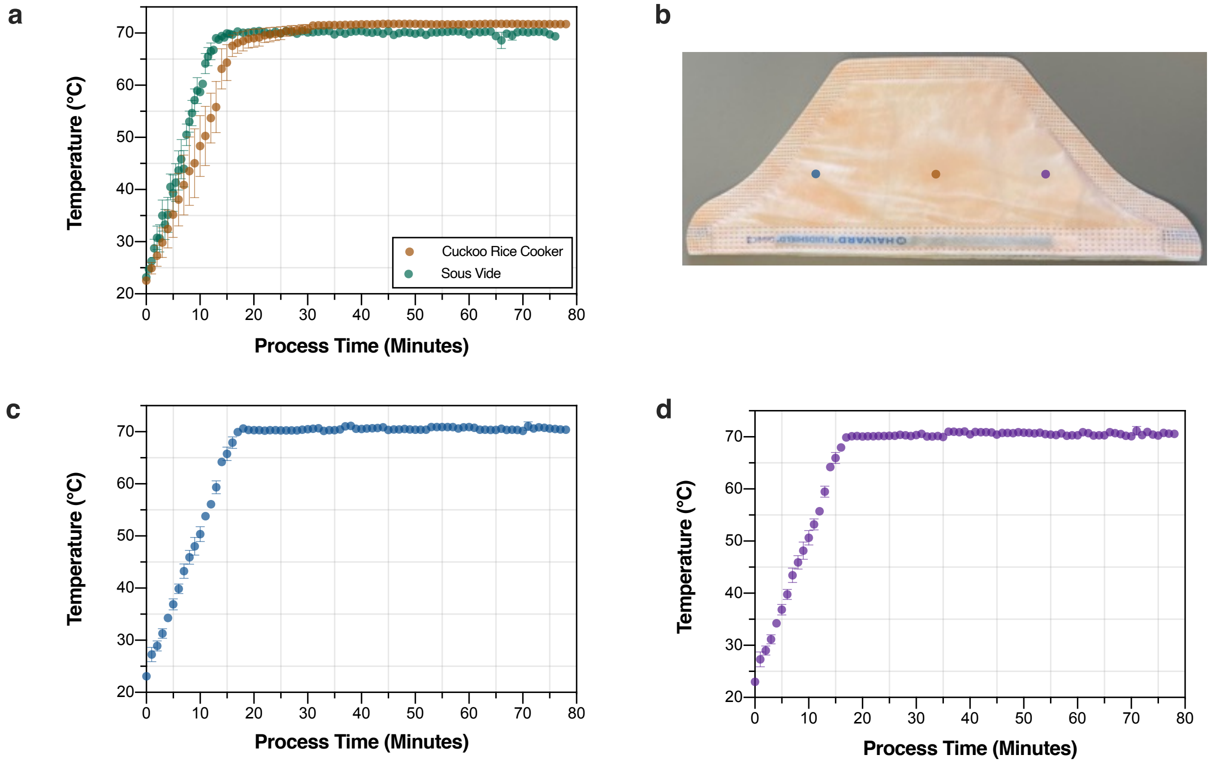


**Fig S1. Average temperature profile of FFRs over one round of treatment. (a)** Temperature was recorded for the duration of the treatment using a wireless sensor (Inkbird Thermometer IBS-TH1 for the rice cooker and MeatStick Wireless Thermometer 4335995327 for the sous vide machine). **(b)** The temperature was measured at two lateral locations of a control mask (blue and purple dots) in addition to the medial location (orange dot) previously used in **(a)**. **(c** and **d)** Measurements at each lateral position was recorded for the duration of one round of treatment and the average temperature profiles over three replicate measurements are shown.

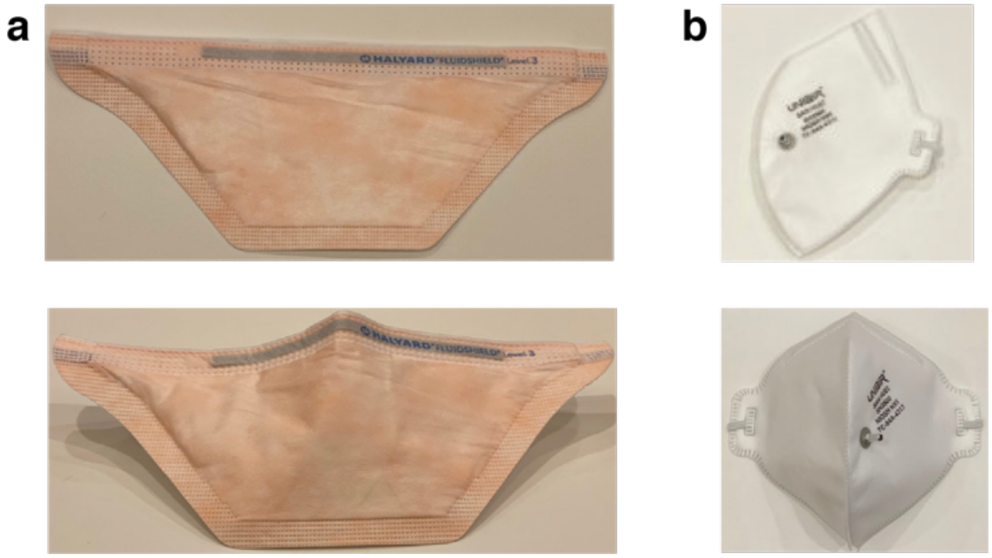


**Fig S2. FFR models. (a)** Halyard Fluidshield 3 FFR. **(b)** Uniair San Huei FFR. Port for quantitative fit testing visible in the photo.

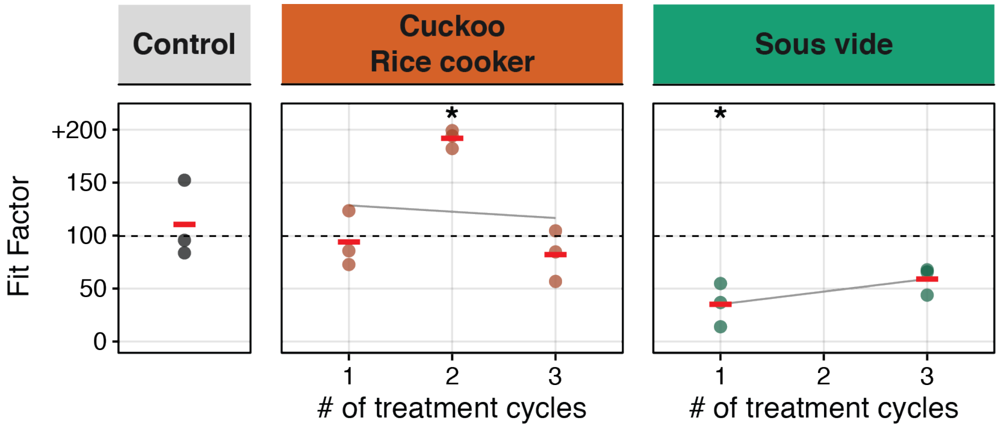


**Fig S3. Average Uniair San Huei FFR fit factor before and after treatment.** Overall fit factor as a function of treatment cycles for each condition: control (left), using a rice cooker (center) and a sous vide machine (right). Fit factor measures were repeated three times for each FFR (individual dots) and average fit factor is reported to the nearest integer (red bar). Grey regression lines quantify fit factor as a function of the number of treatment cycles (Table S4). Asterisks indicate significant differences in fit between treatment group and control (ANOVA, $p < 0.05$).

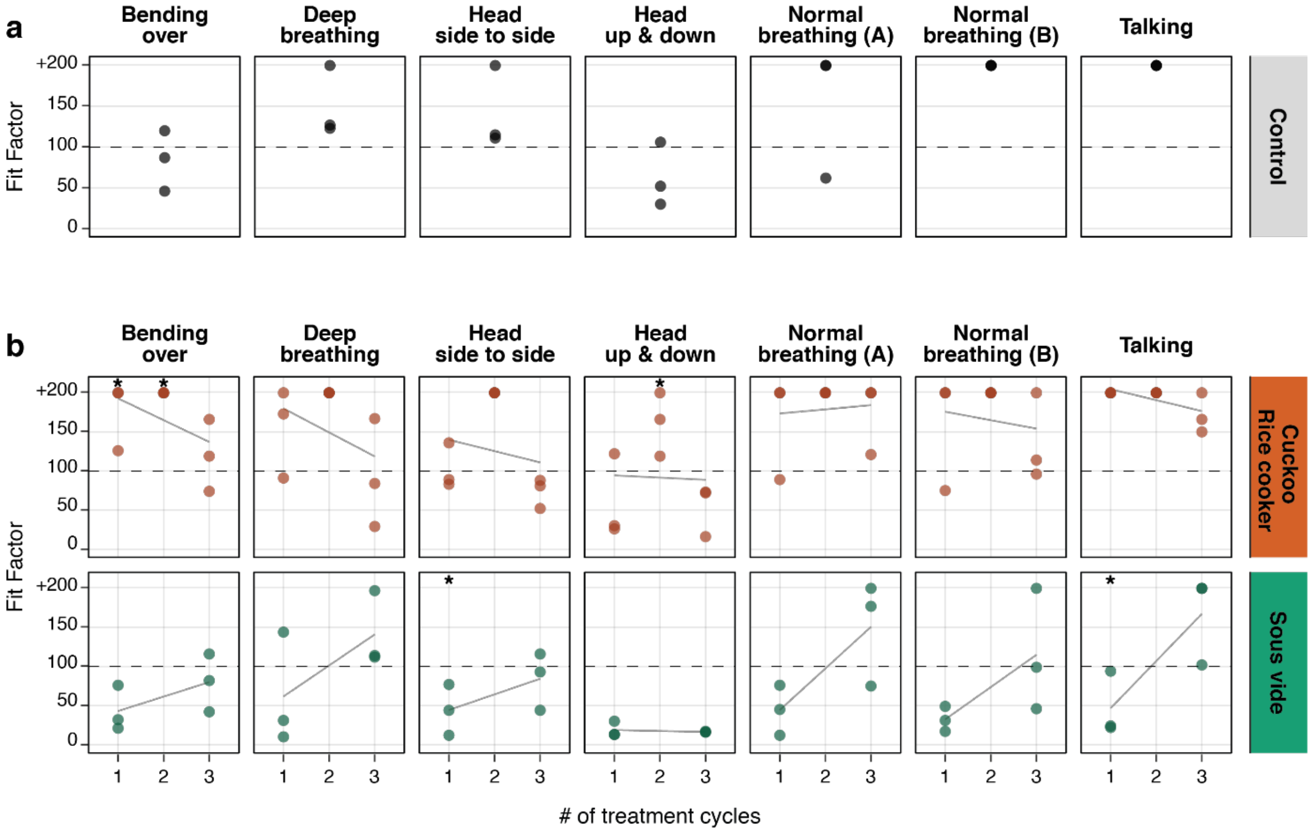


**Fig S4. Uniair San Huei FFR fit for seven standardized testing conditions.** The fit of control **(a)** and heat-treated **(b)** FFRs was assessed three times in each of seven testing conditions (individual dots) for each heat-treatment and control group. Grey lines in (b) signify linear regression fit of fit factor over treatment cycle (Table S5). Asterisks indicate significant differences in mean fit between treatment group and control (ANOVA, $p < 0.05$).

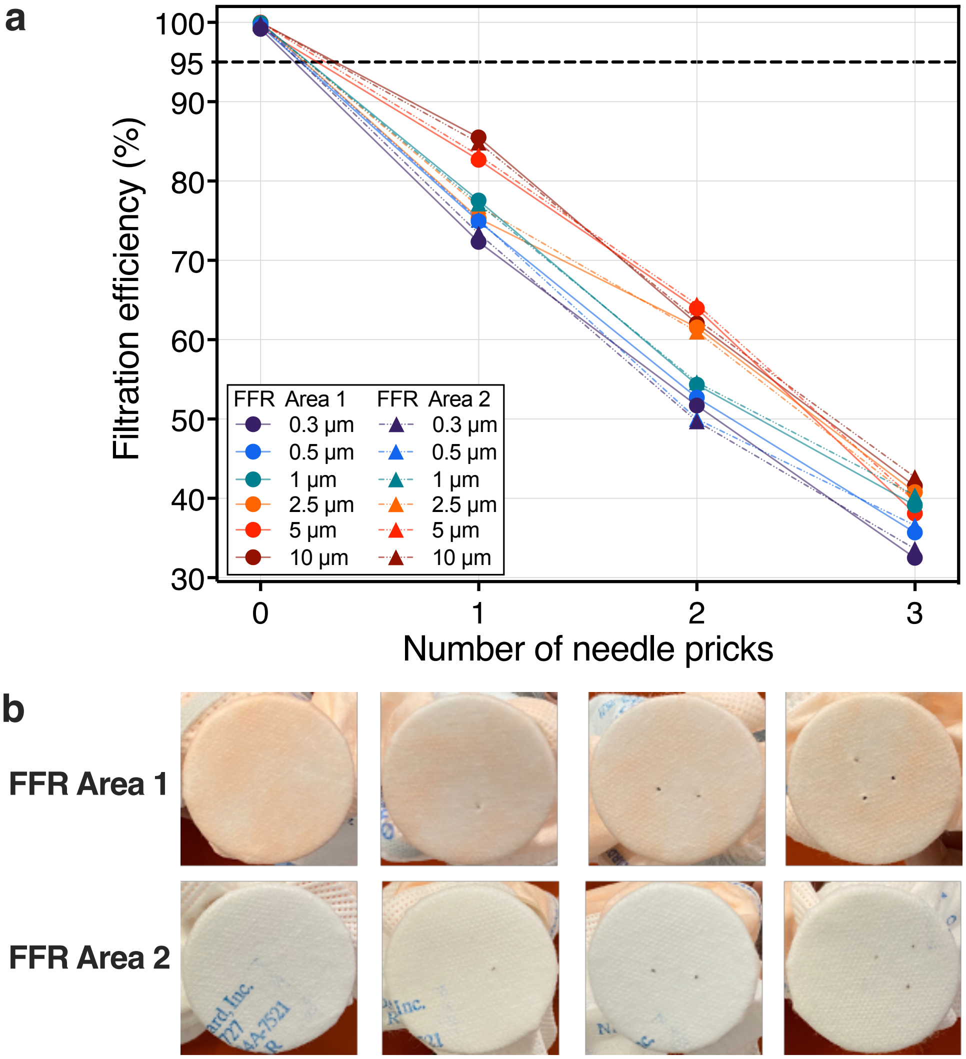


**Fig S5. Filtration efficiency for control and punctured Halyard Fluidshield 3 FFRs. (a)** The filtration efficiency of two N95 FFR areas for different particle sizes (bins of 0.3 μm, 0.5 μm, 1 μm, 2.5 μm, 5 μm, and 10 μm size classes) were determined for an untreated FFR before and after puncturing with an 18-gauge needle once, twice, and three times. Any number of needle pricks decreased the filtration efficiency below the 95% requirement. **(b)** The front view of two FFR regions subject to punctures, fitted to the sample holder, and tested for filtration efficiency is shown.

### SUPPLEMENTARY TABLES S1 – S10

| Virus | Medium | Model | Method | Temp (ºC) | Humidity (%RH) | Time (min) | log reduction in virus | Filtration test passed after # treatments | Fit test passed after # treatments | Source |
|---|---|---|---|---|---|---|---|---|---|---|
| SARS-CoV-2 | N/A | 3M 1860S, 8110S, 8210S, 9105S | Dry heat | 70 | 50 | 60 | > 3 | 10 | 5 | 1 |
| | | AO safety N9504C | | | | 30 | Insufficient | N/A | N/A | |
| | | | | | | 60 | > 3.3 | N/A | N/A | |
| | Bovine serum albumin, tryptone, mucin | 3M 1860, 3M 1870, 3M Vflex 1804, AO Safety 1054 | Steam | 121 | N/A | 15 | > 4.6-5.6 | 1-10 | 4 | 2 |
| | DMEM | AOSafety N9504C | Dry heat | 70 | N/A | 60 | > 3 | 2 | N/A | 3 |
| Murine coronavirus MHV | DMEM | 3M 1860 | Dry heat | 72 | 1 | 30 | Insufficient | N/A | N/A | 4 |
| | | | | 82 | 25 | | | | | |
| | | | | 72 | 25 | | > 3.5 | | | |
| | | 3M 8210 | Humid heat | 75 | 90 | 30 | > 6 | N/A | 10 | 5 |
| H5N1 | Aerosolized allantonic fluid | 3M 1860S, 3M 1870 | Humid heat | 65 | N/A | 30 | > 4.62-4.65 | N/A | N/A | 6 |

**Table S1. Effect of heat and humidity on enveloped viruses on N95 FFRs.** Log viral reduction >3 is sufficient to consider the virus inactivated. N/A: Not Applicable, the parameter was not tested. DMEM refers to Dulbecco's Modified Eagle Medium.

| Type | Appliance | Temperature (°C) | | |
|---|---|---|---|---|
| | | Min | Mean | Max |
| Rice cooker | Cuckoo (CR-0655F) | 69.7 | 71.7 | 72.2 |
| | Aroma (ARC-743-1NGB) | 63.5 | 67.5 | 69.5 |
| Sous vide | AuAg Immersion Circulator (A808, 950 W) | 68.5 | 70.1 | 70.5 |

**Table S2. Temperature maintained after initial heating period in the appliances tested.** The Cuckoo (CR-0655F) *Keep Warm* setting maintained an average temperature of 71.7°C (-2.0°C /+0.5°C). The sous vide AuAg Immersion Circulator (A808, 950 W) has an average temperature of 69.8°C (-1.6°C /+0.4°C). The Aroma (ARC-743-1NGB) *Keep Warm* setting reached an average temperature of 67.5°C (-4.0°C /+2.0°C).

| Rounds of treatment | Treatment | Fit Factor | | | Average Fit Factor | p-value | F statistic | Regression line and $r^2$ |
|---|---|---|---|---|---|---|---|---|
| | | Technical replicate 1 | Technical replicate 2 | Technical replicate 3 | | | | |
| 0 | Control | 108 | 184 | 94 | 129 | - | - | - |
| 1 | Boil | 51 | 141 | 105 | 99 | 0.02 | 12.54 | $y$ = 131–31$x$, $r^2$ = 0.99 |
| 2 | | 54 | 86 | 73 | 71 | 0.00 | 123.67 | |
| 3 | | 33 | 55 | 23 | 37 | 0.00 | 196.67 | |
| 1 | Cuckoo Rice cooker | +200 | 167 | 175 | 181 | 0.31 | 1.32 | $y$ = 187–4.1$x$, $r^2$ = 0.67 |
| 2 | | 167 | 192 | 187 | 182 | 0.28 | 1.53 | |
| 3 | | 165 | +200 | 154 | 173 | 0.24 | 1.93 | |
| 4 | | 184 | 173 | 176 | 178 | 0.08 | 5.70 | |
| 5 | | 144 | 143 | +200 | 162 | 0.18 | 2.57 | |
| 1 | Sous vide | 129 | +200 | 188 | 172 | 0.39 | 0.91 | $y$ = 206–22.8$x$, $r^2$ = 0.66 |
| 2 | | 49 | 186 | 199 | 145 | 0.37 | 1.04 | |
| 3 | | +200 | +200 | 137 | 179 | 0.53 | 0.47 | |
| 4 | | 78 | 98 | 193 | 123 | 0.12 | 3.89 | |
| 5 | | 72 | 76 | 59 | 69 | 0.00 | 36.76 | |

**Table S3. Average Halyard Fluidshield 3 FFR fit factor and technical replicates.** After exposing FFRs to several methods and rounds of treatment, the fit factor was estimated with the PortaCountPro+. The fit factor was estimated three times for each FFRs and the average fit factor is reported to the nearest integer. Control FFR was not subjected to any treatment. Analysis of variance p-values shown for variation in means among treatment group and control. Values rounded to two decimal places. Grey regression line overlaid for each treatment method and category (y ~ mx + b) in Fig 2.

| Rounds of treatment | Treatment | Fit Factor | | | Average Fit Factor | p-value | F statistic | Regression line and $r^2$ (x=reps) |
|---|---|---|---|---|---|---|---|---|
| | | Technical replicate 1 | Technical replicate 2 | Technical replicate 3 | | | | |
| 0 | Control | 96 | 84 | 153 | 111 | - | - | - |
| 1 | Cuckoo Rice cooker | 73 | 86 | 124 | 94.3 | 0.56 | 0.4 | $y$ = 135–6$x$, $r^2$ = 0.01 |
| 2 | | 183 | +200 | 195 | 192.6 | 0.02 | 13.94 | |
| 3 | | 85 | 105 | 57 | 82.3 | 0.32 | 1.27 | |
| 1 | Sous vide | 55 | 37 | 14 | 35.3 | 0.04 | 9.64 | $y$ = 23.3–12$x$, |
| 2 | | - | - | - | - | - | - | |
| 3 | | 73 | 86 | 124 | 59.3 | 0.08 | 5.21 | |

**Table S4. Average FFR fit factor and technical replicates Uniair San Huei**
After exposing FFRs to several methods and rounds of treatment, the fit factor was estimated with the PortaCountPro+. The fit factor was estimated three times for each FFRs and the average fit factor is reported to the nearest integer. The control FFR was not subjected to any round of treatment. Analysis of variance p-values shown for variation in means among treatment group and control. Values rounded to two decimal places. Grey regression line overlaid for each treatment method and category (y ~ mx + b) in Fig S3.

| Treatment | Category | Rounds of treatment | p-value | F statistics | Regression line and $r^2$, (x=reps) |
|---|---|---|---|---|---|
| Cuckoo Rice cooker | Bending over | 1 | 0.05 | 7.76 | $y = 221–28.8x$, $r^2 = 0.26$ |
| | | 2 | 0.01 | 29.20 | |
| | | 3 | 0.36 | 1.07 | |
| | Deep breathing | 1 | 0.92 | 0.01 | $y = 211-30.7x$, $r^2 = 0.17$ |
| | | 2 | 0.12 | 3.99 | |
| | | 3 | 0.30 | 1.44 | |
| | Head side to side | 1 | 0.31 | 1.38 | $y = 154-14.5x$, $r^2 = 0.04$ |
| | | 2 | 0.12 | 3.99 | |
| | | 3 | 0.09 | 4.84 | |
| | Head up & down | 1 | 0.94 | 0.01 | $y = 97.2-2.83x$, $r^2 = 0.001$ |
| | | 2 | 0.04 | 9.23 | |
| | | 3 | 0.77 | 0.09 | |
| | Normal breathing (A) | 1 | 0.89 | 0.02 | $y = 168+5.33x$, $r^2 = 0.01$ |
| | | 2 | 0.37 | 1.00 | |
| | | 3 | 0.73 | 0.14 | |
| | Normal breathing (B) | 1 | 0.37 | 1.00 | $y = 187–10.8x$, $r^2 = 0.03$ |
| | | 2 | 0.37 | 1.00 | |
| | | 3 | 0.12 | 3.89 | |
| | Talking | 1 | 0.37 | 1.00 | $y = 219–14x$, $r^2 = 0.41$ |
| | | 2 | 0.37 | 1.00 | |
| | | 3 | 0.13 | 3.61 | |
| Sous vide | Bending over | 1 | 0.20 | 2.31 | $y = 24.5+18.5x$, $r^2 = 0.32$ |
| | | 3 | 0.89 | 0.02 | |
| | Deep breathing | 1 | 0.14 | 3.31 | $y = 22+39.7x$, $r^2= 0.39$ |
| | | 3 | 0.82 | 0.06 | |
| | Head side to side | 1 | 0.05 | 7.98 | $y = 24.3–20x$, $r^2 = 0.33$ |
| | | 3 | 0.18 | 2.57 | |
| | Head up & down | 1 | 0.13 | 3.57 | $y = 19.8–1.17x$, $r^2 = 0.04$ |
| | | 3 | 0.11 | 4.21 | |
| | Normal breathing (A) | 1 | 0.09 | 4.89 | $y = -8.83+53.2x$, $r^2 = 0.61$ |
| | | 3 | 0.96 | 0.00 | |
| | Normal breathing (B) | 1 | 0.00 | 327.65 | $y = -9+41.3x$, $r^2 = 0.45$ |
| | | 3 | 0.13 | 3.54 | |
| | Talking | 1 | 0.00 | 41.95 | $y = -13.7+60.3x$, $r^2 = 0.69$ |
| | | 3 | 0.37 | 1.00 | |

**Table S5. Analysis of variance among treatment category groups and control Uniair San Huei.** F- and p-values denote the variation in means among treatment category groups and control in an analysis of variance. Grey regression line overlaid for each treatment method and category (y ~ mx + b) in Fig S4.

| Rounds of treatment | Treatment | Technical replicate | Bending over | Deep breathing | Head side to side | Head up & down | Normal breathing (A) | Normal breathing (B) | Talking |
|---|---|---|---|---|---|---|---|---|---|
| 0 | Control | 1 | 52 | 116 | 111 | 126 | 178 | 123 | 181 |
| | | 2 | 122 | +200 | +200 | +200 | +200 | +200 | +200 |
| | | 3 | 57 | 127 | 119 | 81 | 112 | 111 | 99 |
| 1 | Boil | 1 | 63 | 28 | 42 | 98 | 42 | 87 | 63 |
| | | 2 | 127 | 118 | 128 | 146 | 163 | 172 | 154 |
| | | 3 | 99 | 108 | 96 | 120 | 79 | 124 | 128 |
| 2 | | 1 | 51 | 52 | 53 | 48 | 71 | 88 | 42 |
| | | 2 | 73 | 116 | 63 | 65 | 180 | 110 | 78 |
| | | 3 | 48 | 55 | 72 | 84 | 49 | +200 | 178 |
| 3 | | 1 | 61 | 20 | 32 | 39 | 26 | 72 | 26 |
| | | 2 | 51 | 64 | 69 | 47 | 55 | 52 | 52 |
| | | 3 | 27 | 13 | 30 | 18 | 46 | 20 | 35 |
| 1 | Cuckoo Rice cooker | 1 | +200 | +200 | +200 | +200 | +200 | +200 | +200 |
| | | 2 | +200 | +200 | 83 | +200 | +200 | +200 | +200 |
| | | 3 | 150 | 119 | +200 | +200 | +200 | +200 | +200 |
| 2 | | 1 | +200 | +200 | +200 | +200 | 133 | +200 | +200 |
| | | 2 | +200 | +200 | +200 | +200 | +200 | +200 | 154 |
| | | 3 | +200 | +200 | +200 | 159 | +200 | 95 | +200 |
| 3 | | 1 | 168 | 167 | +200 | +200 | 101 | 183 | +200 |
| | | 2 | +200 | +200 | +200 | +200 | +200 | +200 | +200 |
| | | 3 | 65 | +200 | +200 | +200 | +200 | +200 | +200 |
| 4 | | 1 | 133 | +200 | +200 | +200 | +200 | 177 | +200 |
| | | 2 | 124 | 185 | 148 | 185 | +200 | +200 | +200 |
| | | 3 | 195 | +200 | +200 | +200 | 133 | 138 | +200 |
| 5 | | 1 | 74 | +200 | 183 | +200 | +200 | 121 | 162 |
| | | 2 | 100 | 136 | +200 | 140 | 109 | +200 | +200 |
| | | 3 | +200 | +200 | +200 | +200 | +200 | +200 | +200 |
| 1 | Sous vide | 1 | 188 | 153 | +200 | 44 | +200 | +200 | +200 |
| | | 2 | +200 | +200 | +200 | +200 | +200 | +200 | +200 |
| | | 3 | 148 | +200 | 181 | +200 | +200 | +200 | +200 |
| 2 | | 1 | 27 | 56 | 47 | 56 | 68 | 59 | 63 |
| | | 2 | 136 | +200 | +200 | +200 | 190 | +200 | +200 |
| | | 3 | 190 | +200 | +200 | +200 | +200 | +200 | +200 |
| 3 | | 1 | +200 | +200 | +200 | +200 | 198 | +200 | +200 |

| | | | | | | | | |
|---|---|---|---|---|---|---|---|---|
| | 2 | +200 | +200 | +200 | +200 | +200 | +200 | +200 |
| | 3 | +200 | +200 | 78 | +200 | +200 | 75 | +200 |
| | 1 | 32 | 110 | 99 | 72 | 63 | 46 | 114 |
| 4 | 2 | 76 | 116 | 99 | 92 | 108 | 99 | 110 |
| | 3 | 159 | +200 | +200 | +200 | +200 | 197 | +200 |
| | 1 | 68 | 45 | 77 | 123 | 99 | 199 | 43 |
| 5 | 2 | 37 | 135 | 56 | 77 | 133 | 110 | 108 |
| | 3 | 57 | 62 | 51 | 57 | 71 | 59 | 63 |

**Table S6. Halyard Fluidshield 3 FFR fit for different testing categories.** After exposing FFRs to several methods and rounds of treatment, the fit for different testing categories was estimated with the PortaCountPro+. The fit was estimated three times for each category. Control FFRs was not subjected to any round of treatment.

| Treatment | Bending over | Deep breathing | Head side to side | Head up & down | Normal breathing (A) | Normal breathing (B) | Talking |
|---|---|---|---|---|---|---|---|
| **Boil** | $y = 117–25x$<br>$r^2 = 0.52$ | $y = 110–26.2x$<br>$r^2 = 0.30$ | $y = 110–22.5x$<br>$r^2 = 0.38$ | $y = 161–43.3x$<br>$r^2 = 0.82$ | $y = 131–26.2x$<br>$r^2 = 0.17$ | $y = 182–39.8x$<br>$r^2 = 0.37$ | $y = 161–38.7x$<br>$r^2 = 0.36$ |
| **Cuckoo Rice cooker** | $y = 212–16.8x$<br>$r^2 = 0.24$ | $y = 186+0.6x$<br>$r^2 = 0.001$ | $y = 174+4.9x$<br>$r^2 = 0.05$ | $y = 203–3.2x$<br>$r^2 = 0.06$ | $y = 198–6.13x$<br>$r^2 = 0.06$ | $y = 196–4.7x$<br>$r^2 = 0.04$ | $y = 198–1.03x$<br>$r^2 = 0.01$ |
| **Sous vide** | $y = 212–27.9x$<br>$r^2 = 0.34$ | $y = 218–21.9x$<br>$r^2 = 0.28$ | $y = 224–28.3x$<br>$r^2 = 0.38$ | $y = 189–15.7x$<br>$r^2 = 0.02$ | $y = 224–22.9x$<br>$r^2 = 0.34$ | $y = 209–19.6x$<br>$r^2 = 0.19$ | $y = 236–27.3x$<br>$r^2 = 0.41$ |

**Table S7. Regression line and $r^2$: Halyard Fluidshield 3 FFR fit for different testing categories.** Grey regression line overlaid for each treatment method and category (y ~ mx + b) in Fig 2.

| Treatment | Category | Rounds of treatment | p-value | F statistics |
|---|---|---|---|---|
| Boil | Bending over | 1 | 0.00 | 31.93 |
| | | 2 | 0.00 | 332.30 |
| | | 3 | 0.00 | 234.99 |
| | Deep breathing | 1 | 0.02 | 16.68 |
| | | 2 | 0.00 | 36.90 |
| | | 3 | 0.00 | 111.65 |
| | Head side to side | 1 | 0.01 | 20.04 |
| | | 2 | 0.00 | 635.47 |
| | | 3 | 0.00 | 153.94 |
| | Head up & down | 1 | 0.00 | 32.98 |
| | | 2 | 0.00 | 169.38 |
| | | 3 | 0.00 | 369.88 |
| | Normal breathing (A) | 1 | 0.04 | 8.82 |
| | | 2 | 0.07 | 6.22 |
| | | 3 | 0.00 | 342.68 |
| | Normal breathing (B) | 1 | 0.04 | 8.88 |
| | | 2 | 0.12 | 3.98 |
| | | 3 | 0.00 | 102.08 |
| | Talking | 1 | 0.22 | 2.17 |
| | | 2 | 0.21 | 2.19 |
| | | 3 | 0.01 | 21.18 |
| Cuckoo Rice cooker | Bending over | 1 | 0.37 | 1.00 |
| | | 2 | 0.37 | 1.00 |
| | | 3 | 0.24 | 1.89 |
| | | 4 | 0.09 | 5.09 |
| | | 5 | 0.12 | 3.85 |
| | Deep breathing | 1 | 0.37 | 1.00 |
| | | 2 | 0.37 | 1.00 |
| | | 3 | 0.37 | 1.00 |
| | | 4 | 0.37 | 1.00 |
| | | 5 | 0.37 | 1.00 |
| | Head side to side | 1 | 0.37 | 1.00 |
| | | 2 | 0.37 | 1.00 |
| | | 3 | 0.37 | 1.00 |
| | | 4 | 0.37 | 1.00 |
| | | 5 | 0.37 | 1.00 |
| | Head up & down | 1 | 0.37 | 1.00 |
| | | 2 | 0.37 | 1.00 |
| | | 3 | 0.37 | 1.00 |
| | | 4 | 0.37 | 1.00 |
| | | 5 | 0.37 | 1.00 |
| | Normal breathing (A) | 1 | 0.37 | 1.00 |
| | | 2 | 0.37 | 1.00 |
| | | 3 | 0.37 | 1.00 |
| | | 4 | 0.37 | 1.00 |
| | | 5 | 0.37 | 1.00 |

| | | | | |
|---|---|---|---|---|
| | Normal breathing (B) | 1 | 0.37 | 1.00 |
| | | 2 | 0.37 | 1.00 |
| | | 3 | 0.37 | 1.00 |
| | | 4 | 0.19 | 2.50 |
| | | 5 | 0.37 | 1.00 |
| | Talking | 1 | 0.37 | 1.00 |
| | | 2 | 0.72 | 0.15 |
| | | 3 | 0.37 | 1.00 |
| | | 4 | 0.37 | 1.00 |
| | | 5 | 0.65 | 0.24 |
| Sous vide | Bending over | 1 | 0.24 | 1.90 |
| | | 2 | 0.16 | 3.02 |
| | | 3 | 0.37 | 1.00 |
| | | 4 | 0.04 | 9.05 |
| | | 5 | 0.00 | 262.46 |
| | Deep breathing | 1 | 0.37 | 1.00 |
| | | 2 | 0.37 | 1.00 |
| | | 3 | 0.37 | 1.00 |
| | | 4 | 0.12 | 3.99 |
| | | 5 | 0.01 | 19.00 |
| | Head side to side | 1 | 0.37 | 1.00 |
| | | 2 | 0.37 | 1.00 |
| | | 3 | 0.37 | 1.00 |
| | | 4 | 0.12 | 4.00 |
| | | 5 | 0.00 | 307.45 |
| | Head up & down | 1 | 0.37 | 1.00 |
| | | 2 | 0.37 | 1.00 |
| | | 3 | 0.37 | 1.00 |
| | | 4 | 0.12 | 3.92 |
| | | 5 | 0.00 | 34.84 |
| | Normal breathing (A) | 1 | 0.37 | 1.00 |
| | | 2 | 0.32 | 1.00 |
| | | 3 | 0.37 | 1.00 |
| | | 4 | 0.13 | 3.59 |
| | | 5 | 0.01 | 31.12 |
| | Normal breathing (B) | 1 | 0.37 | 1.00 |
| | | 2 | 0.37 | 1.00 |
| | | 3 | 0.37 | 1.00 |
| | | 4 | 0.12 | 3.87 |
| | | 5 | 0.13 | 3.67 |
| | Talking | 1 | 0.37 | 1.00 |
| | | 2 | 0.76 | 0.11 |
| | | 3 | 0.37 | 1.00 |
| | | 4 | 0.49 | 0.57 |
| | | 5 | 0.04 | 8.76 |

**Table S8. Analysis of variance among treatment category groups and control Halyard Fluidshield 3 FFRs.** Results of a 2-Way nested ANOVA comparing fit factor in seven test conditions before and after up to five rounds of heat treatment.

| Treatment | Rounds of treatment | Filtration Efficiency (FE) | | | Pressure Drop | |
|---|---|---|---|---|---|---|
| | | Particle Size (μm) | F.E. of FFR Area 1 (%) | F.E. of FFR Area 2 (%) | FFR Area 1 (mmH2O) | FFR Area 2 (mmH2O) |
| Control | 0 | 0.3 | 99.76 | 98.09 | 38.1 | 38.1 |
| | | 0.5 | 99.88 | 98.68 | | |
| | | 1 | 99.94 | 99.35 | | |
| | | 2.5 | 99.98 | 99.85 | | |
| | | 5 | 99.89 | 99.89 | | |
| | | 10 | 99.88 | 100.00 | | |
| Boil | 1 | 0.3 | 99.95 | 99.77 | 33.0 | 30.5 |
| | | 0.5 | 100.00 | 99.78 | | |
| | | 1 | 100.00 | 100.00 | | |
| | | 2.5 | 100.00 | 100.00 | | |
| | | 5 | 100.00 | 100.00 | | |
| | | 10 | 100.00 | 100.00 | | |
| | 2 | 0.3 | 100.00 | 99.64 | 38.1 | 30.5 |
| | | 0.5 | 100.00 | 99.80 | | |
| | | 1 | 100.00 | 99.89 | | |
| | | 2.5 | 100.00 | 99.92 | | |
| | | 5 | 100.00 | 99.61 | | |
| | | 10 | 100.00 | 99.64 | | |
| | 3 | 0.3 | 98.31 | 98.85 | 35.6 | 27.9 |
| | | 0.5 | 98.76 | 98.61 | | |
| | | 1 | 99.45 | 98.98 | | |
| | | 2.5 | 99.65 | 100.00 | | |
| | | 5 | 99.38 | 100.00 | | |
| | | 10 | 99.42 | 100.00 | | |
| Cuckoo Rice Cooker | 1 | 0.3 | 100.00 | 99.79 | 27.9 | 27.9 |
| | | 0.5 | 100.00 | 99.75 | | |
| | | 1 | 100.00 | 99.80 | | |
| | | 2.5 | 100.00 | 100.00 | | |
| | | 5 | 100.00 | 100.00 | | |
| | | 10 | 100.00 | 100.00 | | |
| | 2 | 0.3 | 100.00 | 98.89 | 33.0 | 30.5 |
| | | 0.5 | 100.00 | 98.90 | | |
| | | 1 | 100.00 | 98.83 | | |
| | | 2.5 | 100.00 | 99.08 | | |
| | | 5 | 100.00 | 100.00 | | |
| | | 10 | 100.00 | 100.00 | | |
| | 3 | 0.3 | 100.00 | 99.77 | 35.6 | 30.5 |
| | | 0.5 | 100.00 | 99.89 | | |
| | | 1 | 100.00 | 99.96 | | |
| | | 2.5 | 100.00 | 99.92 | | |
| | | 5 | 100.00 | 99.72 | | |
| | | 10 | 100.00 | 97.44 | | |
| | 4 | 0.3 | 99.52 | 97.16 | 30.5 | 33.0 |
| | | 0.5 | 99.68 | 96.56 | | |
| | | 1 | 99.70 | 97.82 | | |
| | | 2.5 | 99.85 | 98.61 | | |
| | | 5 | 100.00 | 99.10 | | |

| | | | | | | |
|---|---|---|---|---|---|---|
| | | 10 | 100.00 | 99.34 | | |
| | 5 | 0.3 | 97.21 | 96.55 | 25.4 | 27.9 |
| | | 0.5 | 98.79 | 99.36 | | |
| | | 1 | 97.34 | 99.39 | | |
| | | 2.5 | 96.33 | 100.00 | | |
| | | 5 | 100.00 | 100.00 | | |
| | | 10 | 100.00 | 100.00 | | |
| Sous vide | 1 | 0.3 | 99.90 | 97.73 | 38.1 | 30.5 |
| | | 0.5 | 99.98 | 98.73 | | |
| | | 1 | 99.90 | 97.12 | | |
| | | 2.5 | 100.00 | 97.57 | | |
| | | 5 | 100.00 | 100.00 | | |
| | | 10 | 100.00 | 100.00 | | |
| | 2 | 0.3 | 95.43 | 98.48 | 27.9 | 27.9 |
| | | 0.5 | 97.37 | 99.84 | | |
| | | 1 | 97.93 | 99.82 | | |
| | | 2.5 | 98.89 | 98.91 | | |
| | | 5 | 99.46 | 100.00 | | |
| | | 10 | 99.39 | 100.00 | | |
| | 3 | 0.3 | 99.49 | 98.33 | 27.9 | 27.9 |
| | | 0.5 | 99.67 | 99.88 | | |
| | | 1 | 99.86 | 99.91 | | |
| | | 2.5 | 99.98 | 99.90 | | |
| | | 5 | 99.93 | 99.52 | | |
| | | 10 | 99.78 | 100.00 | | |
| | 4 | 0.3 | 96.61 | 96.92 | 25.4 | 25.4 |
| | | 0.5 | 97.71 | 97.62 | | |
| | | 1 | 97.78 | 96.85 | | |
| | | 2.5 | 97.30 | 97.22 | | |
| | | 5 | 100.00 | 99.21 | | |
| | | 10 | 100.00 | 100.00 | | |
| | 5 | 0.3 | 100.00 | 98.79 | 30.5 | 27.9 |
| | | 0.5 | 100.00 | 99.28 | | |
| | | 1 | 100.00 | 99.69 | | |
| | | 2.5 | 100.00 | 99.81 | | |
| | | 5 | 100.00 | 99.93 | | |
| | | 10 | 100.00 | 99.80 | | |

**Table S9. Filtration efficiency for control and treated Halyard Fluidshield 3 FFRs.** None of the treatment methods tested reduces Halyard Fluidshield 3 FFR filtration performance below 95%. The pressure drop across N95 FFR was determined at a flow rate of 10 lpm.

| Number of pinholes | Pressure Drop (mmH2O) | |
|---|---|---|
| | Across FFR Area 1 | Across FFR Area 2 |
| 0 | 30.5 | 27.9 |
| 1 | 12.7 | 15.2 |
| 2 | 7.6 | 5.1 |
| 3 | 2.5 | 2.5 |

**Table S10. Pressure drop across the N95 material for control and punctured Halyard Fluidshield 3 FFRs.** The pressure drop across the N95 FFR material was determined at a flow rate of 10 lpm before and after one, two, and three punctures made with an 18-gauge needle.
